# Using Generative Artificial Intelligence to Classify Primary Progressive Aphasia from Connected Speech

**DOI:** 10.1101/2023.12.22.23300470

**Authors:** Neguine Rezaii, Megan Quimby, Bonnie Wong, Daisy Hochberg, Michael Brickhouse, Alexandra Touroutoglou, Bradford C. Dickerson, Phillip Wolff

**Author notes:** These authors contributed equally as co-senior authors (Bradford C. Dickerson, Phillip Wolff). **Corresponding authors** Neguine Rezaii, MD Brad Dickerson, MD MGH Frontotemporal Disorders Unit, 149 13^th^ Street, Charlestown, MA, USA 02129.

## Abstract

Neurodegenerative dementia syndromes, such as Primary Progressive Aphasias (PPA), have traditionally been diagnosed based in part on verbal and nonverbal cognitive profiles. Debate continues about whether PPA is best subdivided into three variants and also regarding the most distinctive linguistic features for classifying PPA variants. In this study, we harnessed the capabilities of artificial intelligence (AI) and natural language processing (NLP) to first perform unsupervised classification of concise, connected speech samples from 78 PPA patients. Large Language Models discerned three distinct PPA clusters, with 88.5% agreement with independent clinical diagnoses. Patterns of cortical atrophy of three data-driven clusters corresponded to the localization in the clinical diagnostic criteria. We then used NLP to identify linguistic features that best dissociate the three PPA variants. Seventeen features emerged as most valuable for this purpose, including the observation that separating verbs into high and low-frequency types significantly improves classification accuracy. Using these linguistic features derived from the analysis of brief connected speech samples, we developed a classifier that achieved 97.9% accuracy in predicting PPA subtypes and healthy controls. Our findings provide pivotal insights for refining early-stage dementia diagnosis, deepening our understanding of the characteristics of these neurodegenerative phenotypes and the neurobiology of language processing, and enhancing diagnostic evaluation accuracy.

**One sentence summary:** Computational linguistic analyses of naturalistic speech samples can classify the aphasic variant of patients similarly to expert clinicians and identify well-established and novel linguistic features crucial for classification.

## INTRODUCTION

Language is a vital faculty through which we share our thoughts and feelings, build relationships, and pass on collective knowledge. When this faculty falters, challenges arise, as seen in those with Primary Progressive Aphasia (PPA). PPA is a neurological disorder characterized by the gradual erosion of language abilities while initially leaving other cognitive, affective, and sensorimotor functions largely spared (1). The specific characteristics of the condition vary among individuals but generally fall into one of three variants: nonfluent variant PPA (nfvPPA), characterized by agrammatism and effortful, halting speech; semantic variant PPA (svPPA), marked by difficulties in confrontational naming and single-word comprehension; and logopenic variant PPA (lvPPA), distinguished by core deficits in word retrieval and sentence repetition (2). Patients are typically classified after a comprehensive clinical assessment, including a battery of confrontational language tests. Despite the widespread use of these diagnostic constructs, debate continues about the specific features of each subtype and their distinctiveness (3). Some core features, such as agrammatism, are not well-defined or easily quantified (4), which is one reason a sizeable number of patients are classified as “mixed” PPA (5–9). Another criticism concerns the nature of the categories themselves and the extent to which they reflect the natural partitioning of language abnormalities of PPA. In addtion, there is an alternative hypothesis that patients with PPA, like those with post-stroke aphasia, exhibit a multidimensional spectrum of impairments, with only those at the extremes of certain dimensions being categorically distinct (10). Finally, we propose as an additional challenge that the classification of PPA patients into subtypes may depend, at least in part, on the specific types of confrontational tests included in the assessment battery and may not fully reflect linguistic impairments in naturalistic communication.

We performed this study with two major goals. First, we sought to test the hypothesis that the three major PPA variants are natural categories of aphasic subtypes detectable in patients’ naturalistic speech. We pursued an entirely data-driven approach to discovering natural categories of aphasic subtypes using generative artificial intelligence (AI) Large Language Models (LLMs). Rather than determining whether a neural network can be trained to predict the existing categories of PPA—a process that reinforces prespecified diagnostic constructs— we investigate whether the classic PPA variants emerge from natural correlations in the features of people’s speech. Since LLMs can process language at multiple levels of abstraction, from the syntactic to the conceptual, they do not need the linguistic features to be prespecified or otherwise coded. LLMs’ strengths can be adapted to discover similarities and differences in the speech of a sample of patients. Thus, using this form of generative AI, clusters of patients with similar language characteristics can be discovered from naturalistic speech samples without the biases inherent in the selection of tests in an assessment battery, the frequent performance anxiety experienced by patients undergoing confrontational testing, or the lenses through which clinicians interpret results. Once the clusters of similar patients were discovered, we sought to biologically validate them by analyzing each cluster’s regional atrophy pattern measured with MRI scans.

Our second goal was to identify the linguistic features associated with each category of similar PPA patients. Complementing the capabilities of LLMs, we used an automated parser to identify linguistic features that are used less frequently by each of the PPA variants relative to cognitively unimpaired controls. The value of these linguistic features can be tested by determining the degree to which their impairment predicts the different variants of PPA. Beyond revealing linguistic features used to classify patients with PPA, such an analysis might also provide new insights into the nature of the language system and its breakdown in aphasia. Feature-based classification of PPA has been used in prior research. For example, Fraser et al. (2014) used three different classifiers on a large set of lexicosemantic features (11). They found that such features could be employed by a naïve Bayes classifier to classify patients as either svPPA or nfvPPA with 79% accuracy. Similarly, Themistocleous et al. (2021) used a deep neural network on linguistic features derived from picture descriptions to distinguish all three PPA subtypes with 80% accuracy (12). These models highlight how features from a syntactic parser can be effective in determining different kinds of PPA, but also their limitations. Improving the performance of PPA classifiers might require moving beyond the types of features provided in a syntactic parser.

Therefore, we further sought to investigate one specific linguistic feature that has received minimal attention in PPA research, which is the category of verbs. Prior research has shown that verb comprehension predominantly engages prefrontal cortical areas (13–18), congruent with findings of both verb comprehension and production deficits in nfvPPA patients (19–21). However, verb meanings also activate the left temporoparietal junction, encompassing the posterior lateral temporal cortex and inferior parietal lobule (IPL) (22–28). While patients with lvPPA exhibit cortical degeneration in the left temporoparietal junction, verb processing deficits have not been reported in this variant (19). Studies of patients with post-stroke aphasia demonstrate that individuals with left temporoparietal lesions tend to employ high-frequency (mainly “light”) verbs. In contrast, those with prefrontal lesions produce more low-frequency (mostly “heavy”) verbs (29–34). We have reported a similar finding in nfvPPA patients (35). With this background, we also tested whether separating high- and low-frequency verbs would improve the model’s performance in classifying PPA variants, hypothesizing that patients with nfvPPA would use more low-frequency verbs than those with lvPPA and svPPA.

## RESULTS

### Data-driven analysis using LLMs detects the canonical variants of PPA from connected speech

Using data from 78 PPA patients from the Massachusetts General Hospital (MGH) Frontotemporal Disorders Unit Primary Progressive Aphasia Program, we first evaluated whether the three PPA variant classification system has external validity by employing LLMs to uncover implicit divisions in the ways patients speak when describing the Western Aphasia Battery Picnic Scene (WAB-PS) (36). These individuals had been independently classified as having one of the three PPA variant diagnoses after undergoing a comprehensive clinical assessment by a multidisciplinary team of subspecialists, including a structured history obtained from the patient and an informant, comprehensive medical, neuropsychiatric examinations, and neuropsychological and speech-language pathology assessments (see Materials and Methods).

To measure language similarities across PPA patients, we developed a novel computational method. Our approach hinges on assessing language similarities among patients by measuring the degree to which a large text segment facilitates the prediction of sentences in another text segment. This approach is based on the hypothesis that patients with similar neurological conditions would yield more accurate predictions. We used a two-step algorithm to implement this strategy. First, an LLM named T0 (T-Zero) (37) predicted each sentence in a patient’s language sample. This prediction was based on the sentence immediately preceding it, combined with all sentences produced by the other patient. This procedure was repeated ten times. For each sentence, the highest similarity score generated by a second LLM, T5 (T-five) (38), determined the patients’ sentence similarity. We selected T5 for its pre-training in generating semantic similarity scores on a 1-to-5 scale. We calculated the overall similarity between two patients by averaging these scores across all sentences. This process resulted in a 78 x 78 matrix, representing the similarities among the 78 patients studied.

We next analyzed this matrix for similar clusters of patients using AGNES hierarchical clustering. The AGNES algorithm constructs a hierarchy of clusters in a bottom-up manner (39). A dissimilarity matrix was generated using Pearson correlations. Fig 1 shows the solution that resulted from applying hierarchical clustering to the dissimilarity matrix. The label for each participant represents the individual’s PPA variant as independently diagnosed by expert clinicians using the comprehensive assessment (see Materials and Methods). An inspection of the labels indicates that most patients in each cluster belong to the same clinically-diagnosed PPA variant. The clusters generated from this purely data-driven analysis of connected speech samples agreed with the experts’ clinical diagnoses regarding the variant of PPA in 88.5% of cases.

**Fig 1.**
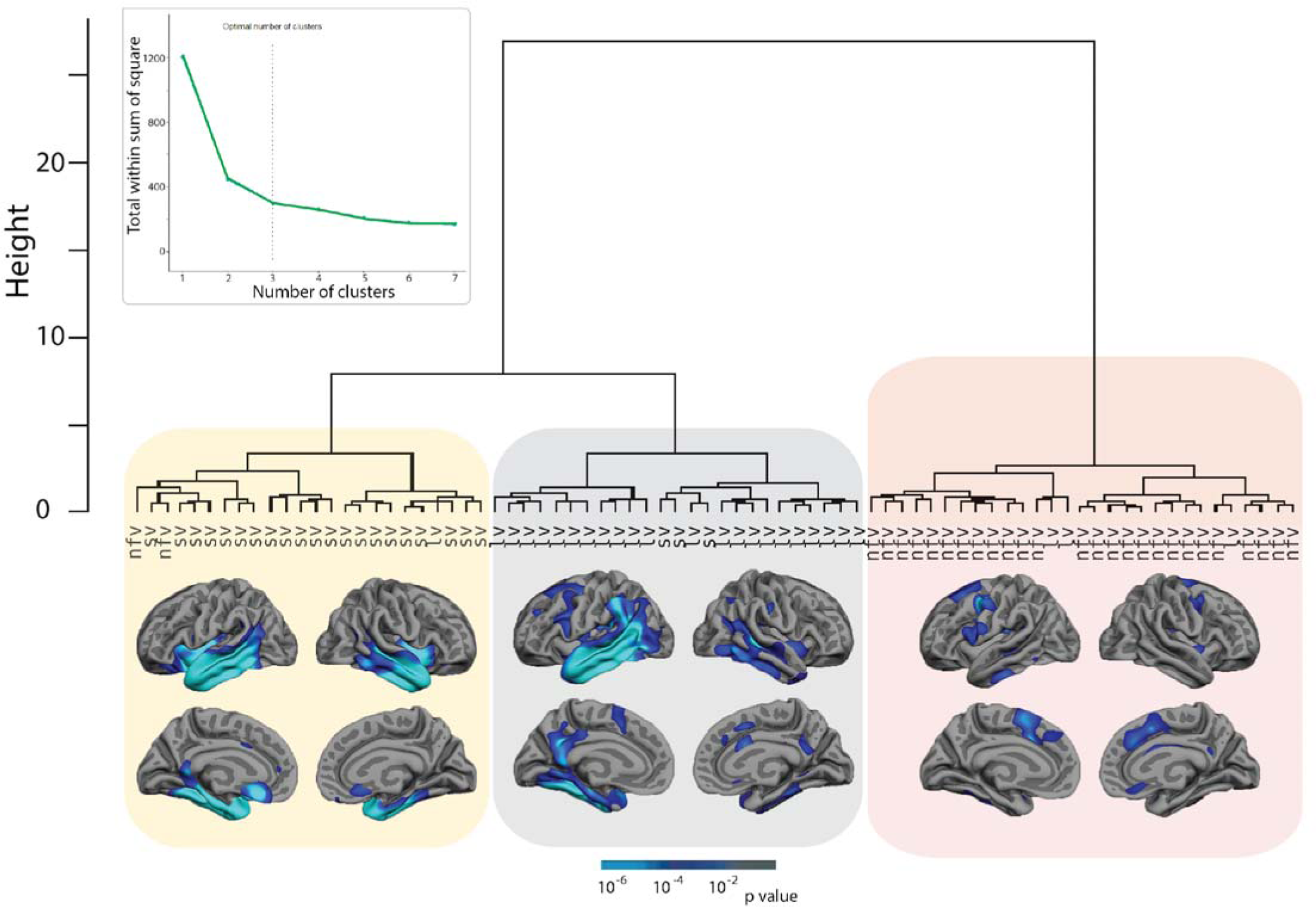
Hierarchical clustering solution of PPA participants’ language samples based on text similarity scores. Participant labels indicate the PPA variant as determined by comprehensive clinical assessments. The first division separates a group largely composed of nfvPPA patients from the remaining patients, while the second division separates a group of mostly svPPA patients from a group of mostly lvPPA patients. The inset shows the total within-cluster sum-of-squares error as a function of the number of clusters, which demonstrates that clusters beyond 3 offer minimal reduction in error, implying that the optimal number of clusters is 3. The total within-cluster sum of squares was based on the dissimilarity matrix used to generate the hierarchical clustering solution. Cortical surface maps indicate areas where each of the three clusters of PPA patients had significantly greater atrophy than age-matched controls, largely recapitulating the atrophy patterns typically seen in each variant. All between-group comparisons are significant at *p* < 0.01.

Participants were partitioned into 1 through 7 clusters from this hierarchical clustering solution. The total within-cluster sum-of-squares error (SSE) for each partition was calculated from the dissimilarity matrix used to generate the hierarchical clusters. The total SSEs for the different cluster numbers are plotted in the inset of Fig 1. The SSEs declined rapidly as the number of clusters increased from 1 to 3, then leveled off, implying a three-cluster solution according to the elbow criterion (40).

We examined the biological validity of the data-driven clusters of PPA patients identified above using an analysis of regional cortical atrophy of each of the three clusters of patients compared with an age-matched group of control participants (Fig 1; see Materials and Methods for details of the analytic approach). Patients in the first cluster, mostly made up of svPPA cases, exhibited prominent bilateral (left more than right) atrophy in the temporal pole, inferior, middle, and superior temporal regions, insula, and orbitofrontal cortex. Patients in the second cluster, largely composed of lvPPA cases, exhibited asymmetrical atrophy in the left anterior temporal cortex, inferior parietal lobule, superior and middle temporal gyri, with weaker effects in dorsolateral prefrontal cortex and inferior frontal gyrus. Patients in the third cluster, largely composed of nfvPPA cases, exhibited atrophy in dorsolateral prefrontal cortex (left more than right), bilateral dorsomedial and mid-cingulate cortex, left inferior frontal gylrus, and left inferior temporal gyrus. These atrophy patterns of clusters of patients segregated by the purely data-driven LLM analysis closely recapitulate the localization of cortical atrophy that is well established for each of these variants using comprehensive clinical evaluations.

### Linguistic features in connected speech distinguish the three PPA variants

We demonstrated above that an unsupervised AI method employing LLMs could differentiate the three major PPA variants from a brief connected speech sample in a manner highly consistent with that of expert clinicians. The findings imply that much of the information needed for classifying variants is present in people’s connected speech. Although successful, this data-driven analysis does not specify the information used by the LLM to compute these similarities. To address this limitation, we used Natural Language Processing (NLP) methods to identify the linguistic features in the connected speech samples that best distinguish the PPA variants. We then performed a clustering analysis on the linguistic features to see whether they provide convergent support for the clusters that emerged from the LLM analysis.

As one hypothesized language feature, we divided verbs into high and low-frequency groups. Since there is no single agreed-upon list of heavy and light verbs, we adopted a quantitative approach to detect a potential natural division in verb distribution using these corpora: The Switchboard Dialog Act Corpus (41), the Santa Barbara Corpus of Spoken American English (42), and the Corpus of Contemporary American English (COCA) (43). The frequencies from the top 966 most frequent verbs from the COCA, which contained the highest number of verbs than other corpora, were converted into a distance matrix by computing the absolute difference in frequency between each verb and every other verb. The resulting distance matrix was submitted to the scikit-learn 1.3.0 k-means clustering (44). Cluster analyses were conducted by setting the number of clusters from 2 to 50. The most frequently occurring cluster contained the first 13 verbs: *be, have, do, go, say, know, get, think, see, come, want, make,* and *take*. Interestingly, these 13 verbs were among the most frequent verbs in the other two corpora (see Supplemental Material 1). Despite being only a small group, these high-frequency verbs made up 56% of the verbs produced by patients and 54% of the verbs produced by healthy subjects in this study. In this analysis, these top 13 verbs were classified as high-frequency, while all other verbs were categorized as low-frequency verbs.

Additional linguistic features were identified using the Stanza NLP toolkit (45). The Stanza parser identified 103 language features. Of these, we retained features with at least three observations for further analysis, resulting in a list of 84 features. These retained features consisted of part-of-speech (N=27, e.g., noun and preposition), syntactic relations expressed in universal dependencies (N=34, e.g., nominal case and clausal complement), and morphosyntactic features (N=23, e.g., past tense and possessive).

To capture language deficits, we reduced the 84 features to those that appeared significantly less frequently than in the control group for at least one of the variants. This was achieved by combining the frequency counts across the patients in the three variants, resulting in a 3 x 84 (group x linguistic feature) matrix. A contingency matrix was constructed by subtracting the expected frequencies from the raw counts and dividing by the square root of the expected value to give standardized residuals with respect to healthy controls (see Materials and Methods). We applied the extended version of Fisher’s exact test to a 3 (groups) x 84 (features) table to determine the probability of the residuals (46). The method indicated that 52 of the original 84 features were associated with significantly negative residuals for at least one of the variants, with family-wise error correction managed by the Simes method (47). As a result, the original matrix was further reduced to a 78 (patient) x 52 (feature) frequency matrix. The counts in the 78 x 52 matrix were compared against expected values based on the healthy controls to generate residuals. We retained only the negative residuals in the 78 x 52 matrix and set the positive residuals to 0. Transposing the matrix allowed us to assess all pairwise dissimilarities with respect to Pearson correlations across participants. The dissimilarity matrix was subsequently analyzed using agglomerative (AGNES) hierarchical clustering with the flexible weight average linking approach, with the par.method set to 0.6 and assuming 1 to 7 clusters. The total within-cluster SSE was calculated for each cluster number and plotted in the inset of Fig 3. The SSEs declined rapidly as the number of clusters increased from 1 to 3, then leveled off, implying that the linguistic features fall into three main clusters. The phylogenetic tree of these features is shown in Fig 3. The linguistic features in the first cluster (yellow) are associated with nouns, either directly or indirectly, via the modification of nouns through determiners, adjectives, or prepositional phrases. Linguistic features in the second cluster are associated with verb phrases, including low-frequency verbs, tense, and possessives (gray). The third cluster (red) is dominated by linguistic features concerning clauses (e.g., mark, TO, xcomp, ccomp). The third cluster also contains linguistic features indicating highly abstract words with austere, template-like semantics, including pronouns (e.g., Acc, Prs, PRP, Nom) and light verbs (e.g., MD, VERB_HighFreq). Figure 3 highlights how the number of cluster-based similarities in the linguistic features matches the number of clusters derived from similarities between participants, providing converging support for a data-driven division of PPA patients into three clusters.

**Fig 2.**
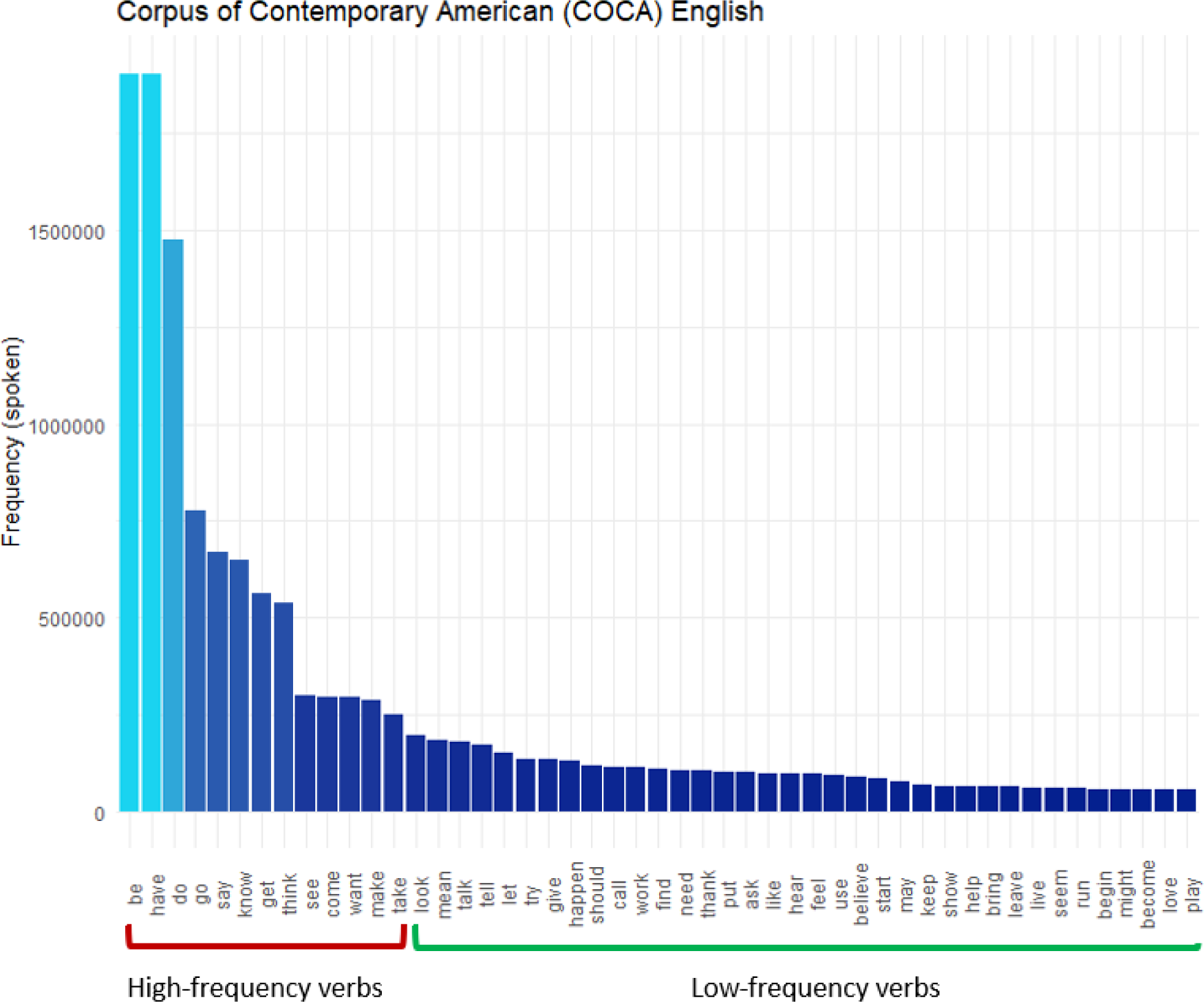
The frequency of the 50 most common verbs in spoken English, according to the Corpus of Contemporary American English, is divided into high-frequency (red) and low-frequency (green) verbs. The frequency of the verb *be* (7,025,941) is so high that it is cut off at the frequency of the second highest verb, *have*.

**Fig 3.**
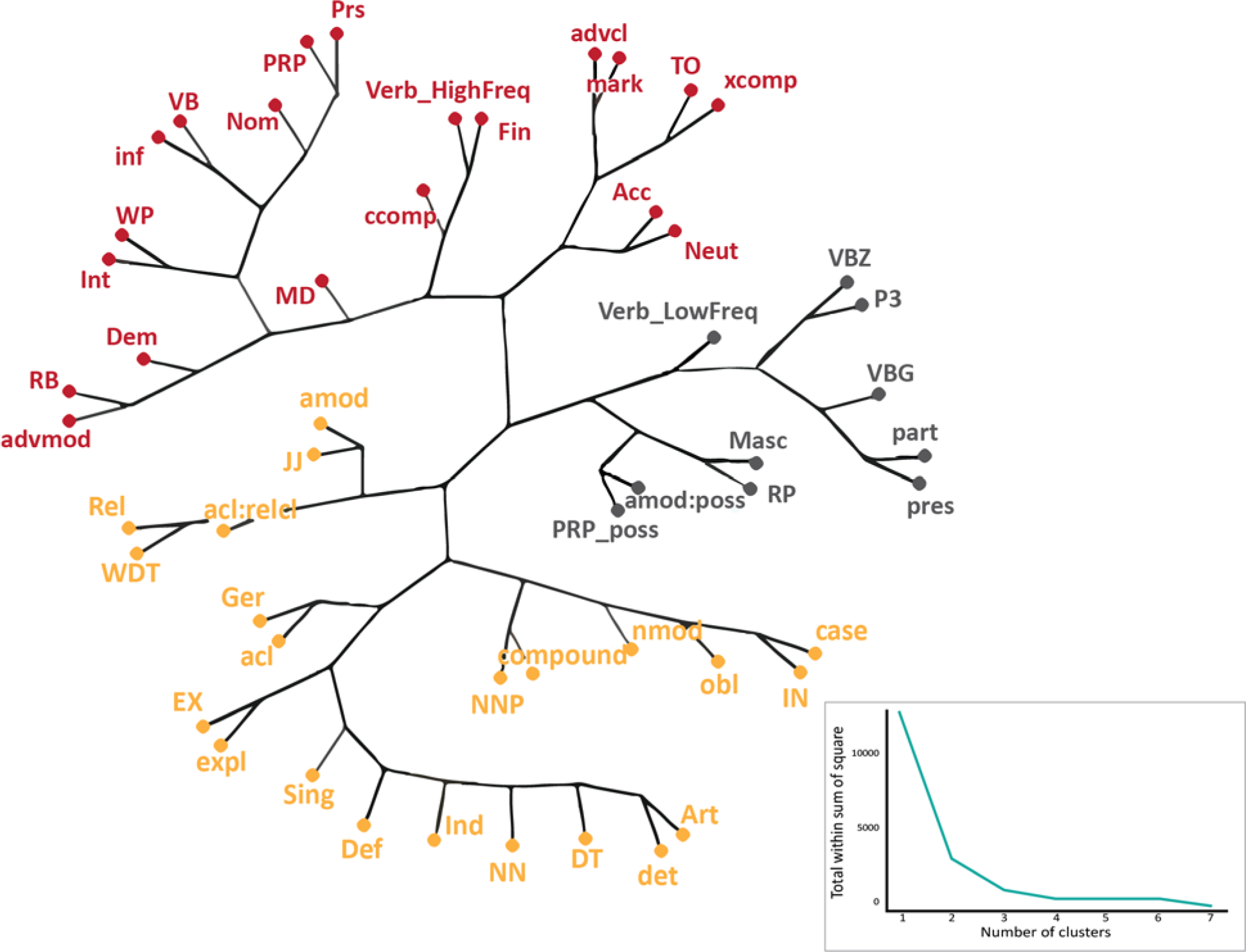
The phylogenetic clustering of the Natural Language Processing-derived linguistic features. The linearized matrix of residuals of features in deficit was submitted to the phylogenetic tree function. The inset shows the scree plot of the total within the sum of squares error for different clusters of the linguistic features based on (dis)similarities used to generate the clusters, indicating three as the optimal number of clusters. Yellow features are associated with nouns, gray features are associated with verb phrases, including heavy verbs, and red features concern clauses and highly abstract words, including pronouns and light verbs. See Supplementary Material 2 for the definition of abbreviated features.

The linguistic feature cluster analysis used features that were in deficit in PPA. However, PPA can also indirectly affect the use of certain language features, leading to increased use of other language features. Fig 4 shows both the positive and negative residuals for the 52 features analyzed in the cluster analysis, providing further insight into the nature of the three clusters. Moving left to right, the first group of linguistic features (nouns) are in greatest deficit for svPPA patients (yellow). The middle group of linguistic features (heavy verbs and verb phrases) are those that are in greatest deficit for lvPPA patients (gray), and the third group of linguistic features (clauses, pronouns, and light verbs) are those in greatest deficit nfvPPA patients (red). In general, the linguistic features associated with the first group center on nouns, as would be expected in svPPA; the linguistic features associated with the second group center on heavy verbs and verb phrases, a novel finding for lvPPA; and the features in the third group center on clauses, pronouns, and light verbs, which are major constituents of syntax as would be expected in nfvPPA.

**Fig 4.**
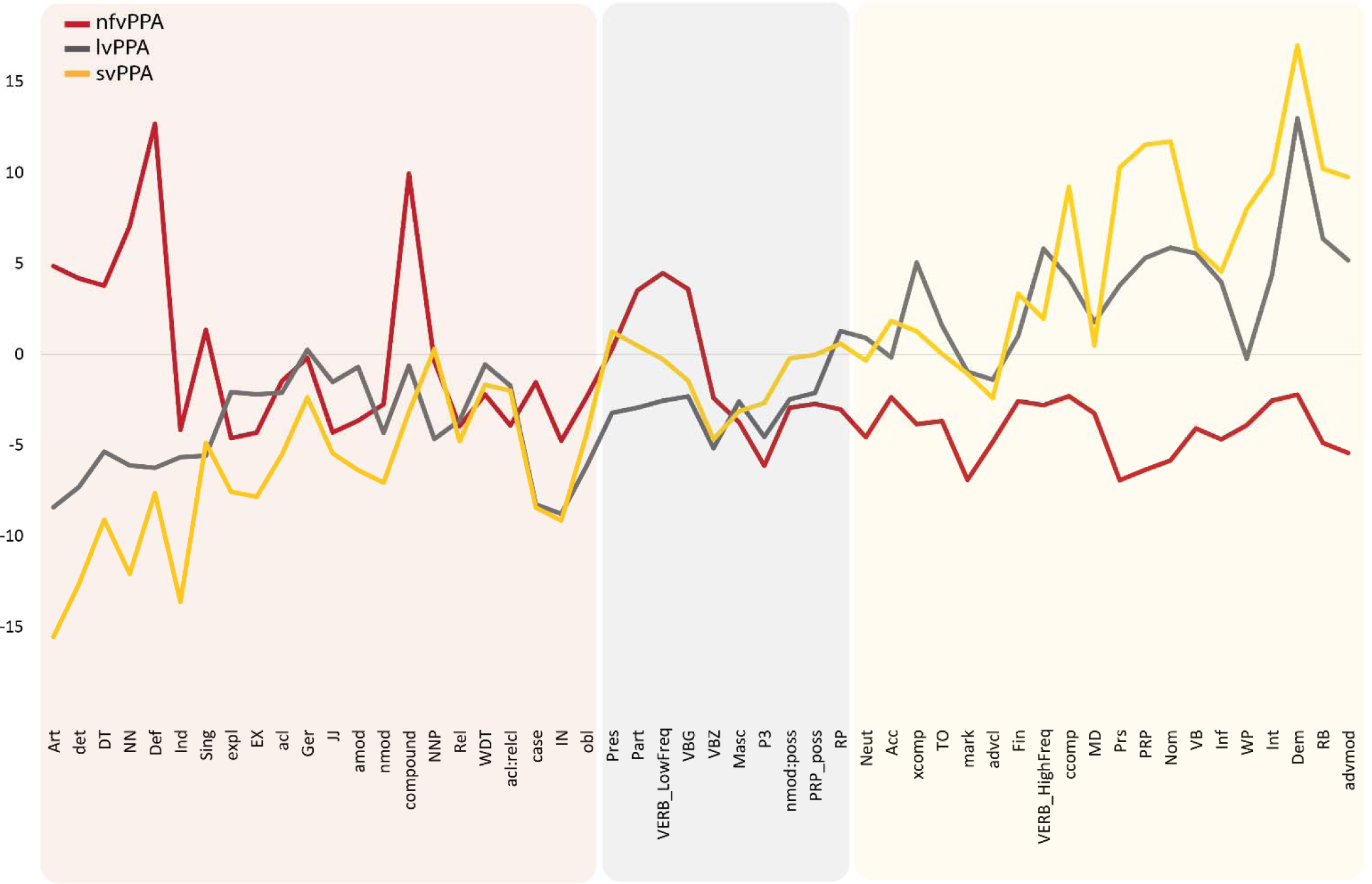
Line chart showing the Natural Language Processing-derived residuals of language features of each PPA variant compared to healthy individuals. Each line shows the residual value of language features for each PPA variant relative to healthy controls. The shaded areas show the features clustered together based on the phylogenetic clustering solution in Fig 2. Features in the shaded yellow region are in most deficit for svPPA patients, the gray shaded region for lvPPA and the red shaded region for nfvPPA. See Supplementary Material 2 for the definition of abbreviated features.

In addition to revealing deficits in the use of linguistic features, the results also show patterns of relative preservation or possibly compensation. Residuals above the 0-line indicate frequency counts that were greater than would be expected from healthy controls. The trade-off between high and low-frequency verbs is shown in Fig 5A, *r*(76)=-0.549, p < 0.001. This pattern helps explain why previous work has not found verbs to be a predictor of PPA variants. Individuals with PPA do not stop using verbs but rather use different types of verbs. Interestingly, if the distinction between high- and low-frequency verbs is disregarded, there was no evidence of a verb deficit across any variant. Standardized (adjusted) Pearson residuals indicated no evidence that verbs as a whole were used more often in nfvPPA individuals than in healthy individuals, r=1.012 *p*=0.32. In lvPPA, there was evidence that verb usage increased relative to healthy controls, r=2.54, *p*=0.012, directly opposite to the deficit of low-frequency specific verbs that we found. Rather, evidence for verb loss only appeared when the verbs were divided into high or low-frequency types. For example, when lvPPA patients have difficulty retrieving the verb *donate,* they may be able to produce *give*. Conversely, patients with nfvPPA produce more low-frequency verbs. A Chi-square test of independence applied to the raw counts indicated that high/low verb frequency was not independent of patient type (nfvPPA, lvPPA), *X*^2^(2)=5.98 *p* < 0.001. Supporting this conclusion, Shan and Gerstenberger’s (2017) extended version of Fisher’s exact test indicated that for nfvPPA individuals, the rate of high-frequency verbs was less than in healthy controls, r=-2.79, *p*=0.006, and the rate of using low-frequency verbs was greater than healthy controls, r=4.47, *p* < 0.001 (46). Further, for lvPPA individuals, the rate of using high-frequency verbs was greater than for healthy controls, r=5.82, *p* < 0.011, and that of low-frequency verbs was marginally less than for healthy controls, r=-2.108, *p*=0.038. This pattern indicates a double dissociation between PPA variant and verb type.

**Fig 5.**
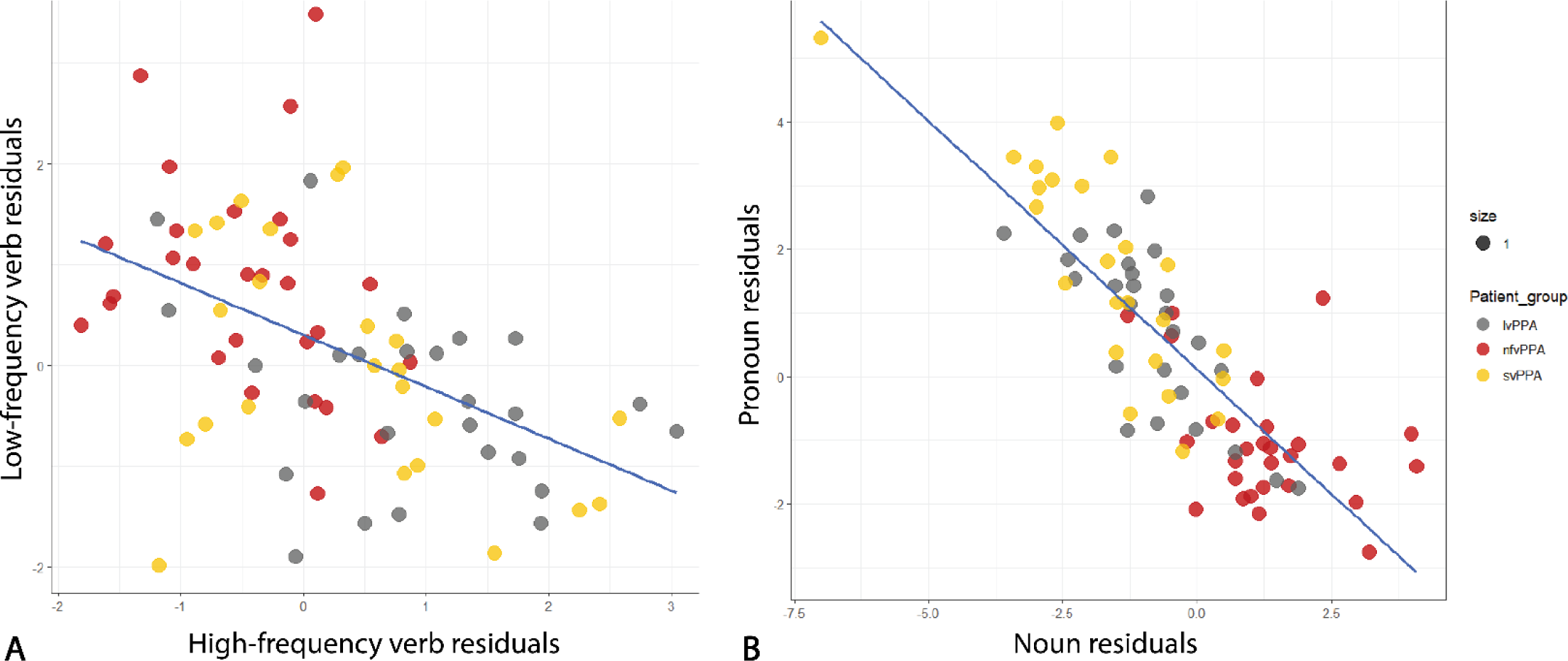
Scatterplots showing trade-offs between low- and high-frequency verbs (A) and nouns and prepositions (B). In panel A, nfvPPA patients tend to occupy the upper left corner, indicating relatively few high-frequency verbs, while lvPPA patients tend to occupy the lower right corner, indicating relatively few low-frequency verbs.

Figure 5B shows a scatter plot indicating a strong negative correlation between pronoun usage (PRP) and noun (NN) usage, *r*(76)=-0.827, *p* < 0.001. Individuals with lvPPA used high-frequency verbs more often than healthy controls, whereas individuals with nfvPPA used low-frequency verbs more often than healthy controls. An analogous pattern of results was observed between different kinds of nouns (common and pronouns) and PPA variant. A Chi-square test of independence applied to the raw counts indicated that noun and pronoun counts were not independent of PPA variant (nfvPPA, svPPA), r=177.9, *p* < 0.001. Fisher’s exact test indicated that for nfvPPA individuals, the rate of using common nouns was higher than in healthy controls, r=6.284, *p* < 0.001, and lower in svPPA individuals, r=-12.51, *p* < 0.001 individuals. Conversely, for nfvPPA individuals, the rate of using pronouns was less than in healthy controls, r=-6.57, *p* < 0.001, while for svPPA individuals, the rate of using pronouns was higher than in healthy controls, r=11.717, *p* < 0.001. The pattern of noun type (common nouns, pronouns) offers yet another example of a double dissociation between PPA variant and language feature.

### NLP-derived linguistic features from connected speech samples robustly predict PPA variant classification

Multinomial logistic regression analysis was conducted to identify which features best differentiated the three PPA variants. Variable selection was achieved by correlating the residuals from the previous analyses with dummy variables representing the participant groups. The resulting correlation coefficients were rank ordered. We used this ordered list of linguistic features to add variables to the model in a forward stepwise manner, with the exception of low-frequency verbs, which were included in the model for theoretical reasons discussed in this work. In addition to low-frequency verbs, the variables included high-frequency verbs, determiners, nouns, pronouns, adverbs, verbs, adverbial modifiers, determiners, clause markers, articles, demonstratives, finites, neuter, nominative case, and personal/possessive pronouns.

The dependent variable of variant contained four categories: nfvPPA, lvPPA, svPPA, and healthy controls. Models were trained on the frequency counts, with healthy controls serving as a reference category. Table 1 shows the logistic coefficient for each predictor, which is the expected amount of change in the logit for each unit change in the predictor. A likelihood ratio test indicated that such a model significantly outperformed a model based on only the constant, *X*2 (51)=270.25, p < 0.001. The 17-variable model after 10-fold cross-validation resulted in an accuracy of 97.88% (sensitivity/recall=0.98; specificity=0.99; precision=0.98). The few errors were evenly distributed across the classes, as shown in the normalized confusion matrix in Supplementary Material 3. When high and low-frequency verbs were combined, the model’s accuracy after 10-fold cross-validation dropped to 74.5% (sensitivity/recall=0.74; specificity=0.90; precision=0.74). A least likelihood test indicated that the model separating high- and low-frequency verbs accounted for more variance than a model that did not distinguish the two kinds of verbs, *X*2 (3)=65.39, *p* < 0.001.

**Table 1.** Coefficient estimates (and standard errors) for the different categories (nvfPP, IvPPA, and svPPA) of the outcome variable.

| Term | nvfPPA |  | lvPPA |  | svPPA |  |
| --- | --- | --- | --- | --- | --- | --- |
|  |  | SE |  | SE |  | SE |
| (Intercept) | 1162.181 | 2901.713 | 981.159 | 3251.751 | 1787.088 | 98.244 |
| VERB_root_HiFrq | -159.465 | 3409.846 | 141.97 | 6764.774 | 56.5 | 1405.275 |
| VERB_root_LoFrq | -85.337 | 6703.728 | -32.426 | 3757.616 | -120.749 | 3555.647 |
| DT | 389.587 | 5645.07 | -206.993 | 8642.115 | 82.722 | 2120.63 |
| NN | 31.411 | 2506.194 | -28.57 | 6668.421 | -36.372 | 2727.295 |
| PRP | -277.162 | 4886.797 | -92.164 | 2069.98 | 37.028 | 7784.414 |
| RB | 91.67 | 3331.921 | -91.858 | 3655.718 | -102.196 | 5296.047 |
| VB | 806.681 | 1970.678 | 796.386 | 6379.979 | 610.893 | 2758.234 |
| advmod | -184.745 | 5586.473 | 56.362 | 8364.208 | 150.403 | 6085.337 |
| det | -418.484 | 3792.571 | 381.71 | 4496.867 | 435.934 | 995.227 |
| mark | -94.698 | 5530.38 | -43.13 | 12620.76 | -130.774 | 1978.209 |
| Art | 51.88 | 5797.921 | -164.897 | 4649.918 | -557.593 | 3991.363 |
| Dem | -174.8 | 5869.075 | 143.721 | 5773.616 | -134.385 | 1758.845 |
| Fin | -104.977 | 5296.543 | -115.268 | 3706.097 | -132.909 | 6687.762 |
| `Inf` | -853.776 | 2278.348 | -763.663 | 7083.154 | -790.04 | 2757.542 |
| Nom | 407.945 | 5310.651 | 115.78 | 4742.165 | 189.788 | 8645.789 |
| Prs | 80.979 | 3754.932 | 49.968 | 4568.466 | 3.088 | 11977.3 |
| Neut | -179.701 | 3737.658 | 55.499 | 4894.679 | 45.816 | 513.784 |

## DISCUSSION

In this study, we leveraged advances in AI and machine learning to classify variants of PPA based on short naturalistic samples of connected speech. Through a data-driven approach, we used LLMs to measure language similarity among pairs of language samples and subsequently applied hierarchical clustering analysis to the resulting similarity matrix. Our analysis revealed the emergence of three main clusters, demonstrating an 88.5% agreement with the independent classification of PPA variants using the international consensus diagnostic criteria. These findings suggest that the three-variant classification of PPA likely reflects natural categorical groups measurable from naturalistic connected speech. Furthermore, as expected based on the predominant clinical variants in each of the three data-driven groups, regional brain atrophy in these groups matched well with the atrophy patterns that are well-established for the three variants. Once we showed that the canonical PPA variants reflect natural kinds, we sought to identify the linguistic features that maximize the distinction between PPA variants. We used an automated syntactic parser to extract the linguistic features most robustly associated with each PPA variant, which sheds new light on dissociable aspects of impaired versus preserved elements of language in the three PPA variants.

We found that patients with svPPA exhibit deficits in nouns as well as linguistic features related to noun modification. These linguistic features include determiners (e.g., DET, Art, WDT, Rel) and features that are used in conjunction with noun phrases, such as expletives (e.g., expl, EX) or other types of noun modifiers (e.g., acl, nmod, compound, JJ, acl:relcl) (See Supplementary Material 2 for the definition of feature abbreviations). SvPPA is primarily associated with cortical atrophy in the anterior temporal lobe region, aligning with reports indicating disruptions in noun processing due to cortical atrophy in the left temporal lobe (13, 16, 48). Patients with lvPPA showed deficits in using low-frequency verbs as well as verb features (e.g., Pres, Part, VBZ, VBG). Patients with lvPPA are also impaired in using features associated with possessive relations (nmod:poss, PRP_poss, Masc in the current dataset). This category also includes particles (e.g., RP), which in this dataset predominantly serve to form phrasal verbs. LvPPA is primarily associated with cortical atrophy in the left posterior lateral temporal and inferior parietal (IPL) regions. Our results align with studies showing that verb meanings recruit the left intraparietal sulcus and inferior parietal lobule (22–24, 49). Finally, patients with nfvPPA exhibit deficits in constructing clauses. This group comprises features related to subordinate clauses (e.g., xcomp, ccomp, mark), clausal modifiers (e.g., advcl), and features frequently involved in subordinate clauses (e.g., WP, VB, Inf, TO, VB). Furthermore, grammatical case relations, which encode the grammatical roles played by noun phrases in sentences, emerge only in the context of clauses (e.g., Nom, Acc). The clause- related features are associated with high-frequency verbs (Verb_HighFreq), which as suggested in the Two-Level Theory of verb meaning (see below), are reflective of basic clausal structures (50–52). Additionally, this third group of features includes pronouns (e.g., PRP, Prs, Nom, Acc, Neut), representing abstract versions of nouns, just as the high-frequency verbs represent abstract versions of low-frequency verbs. NfvPPA is associated with cortical atrophy in the left inferior frontal regions, consistent with studies showing verb impairment in the left prefrontal context (13, 14, 16–18, 53).

Our *a priori* approach to separating high vs. low-frequency verbs in the classification model significantly increased the accuracy from 70% to 98%. In addition to improving the classification model, the high/low-frequency verb distinction could inform the neurolinguistic literature on how the category of verbs is processed across the language network. Inconsistent findings about the neurobiological underpinnings of verbs have been reported in the field. Similar inconsistencies exist in the PPA literature, with some studies reporting verb deficits in nfvPPA while others attributed the deficit to lvPPA, and yet other studies attributing no verb deficit to any PPA variant (see (54) for a review). In this work, we identified a double dissociation between verb frequency and PPA variants, a pattern that could be partially explained by the Two-Level Theory of verb meaning (55–59). This theory suggests that verbs consist of two separate layers of meaning. One layer is the “root” or unique meaning, which captures idiosyncratic semantic features that (a) distinguish each verb in a given class from all the others, (b) are often concrete and modality-specific, and (c) do not interface with grammar (55–59). Another layer is the event structure template, which is (a) common to all the verbs in a given class, (b) composed primarily of schematic predicates and variables for arguments, and (c) relevant to the grammatical properties of all the verbs in a given class. Event structure templates are represented by a limited set of event types such as state, result state, manner, and instrument, which are defined by primitive predicates. In this model, the basic event structure classifies verbs. For instance, the semantic frame [x ACT <INSTRUMENT>] is assigned to instrument verbs like *brush, hammer, saw,* and *shovel*; [x ACT <MANNER>] fits with manner verbs like *jog, run, snore,* and *whistle*. Similarly, other semantic frames can be attributed to state verbs, internally caused state verbs, and change of state verbs. The root of a verb authorizes the event structure based on the meanings it encodes. A root denoting means or manner permits a simple semantic frame, while a result root authorizes a complex frame structure with causing and result subevents, e.g., [x ACT] CAUSE [BECOME [y <SPLIT>]]]. The event structure of a verb influences the spectrum of clausal constructions it can appear in. Essentially, the event structure specifies the syntactic patterns tied to a particular verb meaning (50). Goldberg emphasized the close relation between light verbs and basic clausal structures (51, 52). Specific brain areas linked with light verbs could be those that specify event structure, whereas areas tied to heavy verbs could specify the root of the event structure. Hence, the light verbs used in our study (e.g., *be, do, go*) would likely be linked to certain clausal structures without any particular root connection. Also, heavy verbs might represent solely the root of a verb meaning, a possibility supported by construction grammar (51, 52).

Our findings are consistent with a three-stage model of language production, which posits that language production results from the successive addition of increasingly complex linguistic elements, a concept we refer to as *cumulative complexity*. The model employs time-honored categories of three primary groups of linguistic features: noun-related, verb-related, and clause-related features. This approach implies a sequential order in linguistic processing, commencing with the assembly of noun phrases, followed by the construction of verb phrases, and culminating in the synthesis of a complete sentence. No single brain region is dedicated to semantics or syntax in this model, as previously specified in the classic Wernicke–Lichtheim-Geschwind model. Grammar-related features are distributed throughout the language system. This interpretation is supported by meta-analyses showing that the ordering of cortical processing in the language networks starts in the temporal lobe before moving to the inferior parietal lobule and finally reaching the frontal lobe (60, 61).

Another notable element of the model is that damage to a part of the language networks, e.g., frontoinsular cortex in nfvPPA or temporal pole in svPPA, does not lead simply to a deficit in the production of specific language features relative to healthy individuals, but also to a compensatory increase in other features to attempt to maintain the efficient communication of information. For example, nfvPPA patients who have difficulty using complex syntax (e.g., subordinating clauses) use a higher rate of nouns to all words compared to healthy controls. This finding aligns with our recent work showing that patients with nfvPPA who have difficulty using long and complex structures use more informative words, such as heavy verbs and more content words in their sentences to sustain sentence information (35, 62–64). Similarly, patients with lexicosemantic deficits who have difficulty using nouns relative to other words produce more clause subordination than healthy individuals, as we have recently shown (63). For example, patients with svPPA exhibit a higher rate of embedding and other complex syntactic structures than healthy speakers (48, 63).

Directionality in the language circuit is further supported by the pattern of impairment and potential compensation shown in Fig 4. If the language network is directional, then compensation should be more likely to occur towards the end of the processing pipeline than at the beginning. Fig 4 shows precisely this pattern, with positive residuals emerging more prominently on the right of the pipeline than on the left. Interestingly, however, some compensation appears earlier in the pipeline. Such compensation could be explained by a recursive process by which outputs from the frontal lobe serve as inputs into the temporal lobe, potentially through the ventral stream via the extreme capsule fiber system/longitudinal inferior-frontal-occipital fasciculus or the uncinate fasciculus (27, 65). As proposed by Friederici et al. (2015), the uncinate fasciculus, which connects the frontal operculum and orbitofrontal cortex to the anterior superior temporal gyrus, may be involved in building local syntactic phrases (27).

In summary, this study showcases the efficacy of contemporary generative LLMs in using data-driven analysis of a brief connected-speech sample to categorize patients with Primary Progressive Aphasia (PPA) into one of its three typical variants—a task traditionally accomplished by expert clinicians after exhaustive, specialized assessment typically taking several hours. Leveraging NLP for linguistic feature analysis, this approach identified linguistic features enabling robust classification, including those absent in the current diagnostic criteria, such as the pivotal role of verb categories when segmented into high or low-frequency verbs. Importantly, our methodology has the potential to refine existing diagnostic standards. For instance, the current criteria for subtyping semantic variant PPA (svPPA) and logopenic variant PPA (lvPPA) includes object naming and word retrieval deficiencies, respectively, a differentiation often blurred due to their resemblance, thus creating challenges in clinical practice. However, our findings indicate a more pronounced impairment in noun usage among svPPA patients, while lvPPA patients struggle more with retrieving proper nouns, such as people’s names. Besides offering a robust classification mechanism, our method also unravels insights into the neurobiological mechanisms of verb processing in the language network. The approach in this study can extend to other neurodegenerative conditions, fostering a more objective, theory-neutral categorization system that could provide further insights into the neurobiology of language and other aspects of cognition critical for communication. Future work is needed to test the generalizability of our results on different PPA samples as well as the effectiveness of this method on the classification of PPA patients that do not fit into established variant categories.

## MATERIALS AND METHODS

### Participants

Seventy-eight patients with PPA were recruited from an ongoing longitudinal study at the Primary Progressive Aphasia Program in the Frontotemporal Disorders Unit of Massachusetts General Hospital (MGH). All patients underwent a standard clinical evaluation, comprising a structured history obtained from both patient and informant, comprehensive medical, neurological, and psychiatric history and exams, neuropsychological and speech-language assessments, and a clinical brain MRI scan (66). Ratings on our language scale, called the Progressive Aphasia Severity Scale (PASS), were also included (67). Modeled after the Clinical Dementia Rating Scale (CDR), PASS uses the clinician’s best judgment and integrates information from the patient’s test performance and a companion’s interview. The PASS includes “boxes” for fluency, syntax, word retrieval and expression, repetition, auditory comprehension, single-word comprehension, reading, writing, and functional communication. The PASS Sum-of-Boxes (SoB) is the sum of the box scores. The clinical and demographic information on the patients is shown in Table 2.

**Table 2.** The clinical and demographic information of the participants.

| **Table 2. The clinical and demographic information of the participants** |  |  |  |  |  |  |  |
| --- | --- | --- | --- | --- | --- | --- | --- |
| Group | N | Sex (F:M) | Age | Handedness (right: left) | Years of education | PASS.SOB | MoCA |
| lvPPA | 27 | 10:17 | 68.52 (6.65) | 24:3 | 16.30 (2.40) | 5.34 (2.76) | 21.56 (4.70) |
| nfvPPA | 28 | 16:12 | 68.31 (9.38) | 26:2 | 17.59 (7.23) | 5.58 (3.52) | 20.90 (4.38) |
| svPPA | 23 | 14:9 | 66.44 (8.32) | 20:3 | 16.38 (1.86) | 5.4 (2.30) | 15.56 (7.67) |
| Progressive Aphasia Severity Scale; MoCA: Montreal Cognitive Assessment |  |  |  |  |  |  |  |

In addition, we used data from twenty healthy cognitively unimpaired older adult controls from the Speech and Feeding Disorders Laboratory at the MGH Institute of Health Professions. These individuals had an average age of 65.2 and an average years of education of 15.8. Fifty percent of healthy controls were female, and 75% were right-handed. All study participants provided informed consent in accordance with guidelines established by the Mass General Brigham Healthcare System Institutional Review Boards, which govern human subjects research at MGH and specifically approved this study.

### Language samples

The participants were asked to view a drawing of a family at a picnic from the Western Aphasia Battery– Revised (36) and describe it using as many full sentences as they could. Responses were audio-recorded using an Olympus VN-702PC Voice Recorder in a quiet room and later transcribed into text using the Microsoft Dictate application. The transcriptions were then manually checked for accuracy by a research collaborator blind to patient characteristics.

### LLM data analysis and hierarchical clustering

The analyses used the 11-billion-parameter version of T5, which is available through the Huggingface transformer library (68). The T0 LLM available through the Huggingface transformer library was used to generate guesses about possible sentences (68). Processing was conducted on a Microway Compute Server with four NVIDIA Ampere A100 80GB GPUs. To measure language similarities across PPA patients, we used a two-step algorithm to implement this strategy. Initially, an LLM named T0 (T-Zero) (37) predicted each sentence in a patient’s language sample. This prediction was based on the sentence immediately preceding it, combined with all sentences produced by the other patient. This procedure was repeated ten times. For each sentence, the highest similarity score generated by a second LLM, T5 (T-five) (38), determined the patients’ sentence similarity. We selected T5 for its pre-training in generating semantic similarity scores on a 1-to-5 scale. We calculated the overall similarity between two patients by averaging these scores across all sentences. This process resulted in a 78 x 78 matrix, representing the similarities among the 78 patients studied. The process of discovering clusters was accomplished using hierarchical clustering. The matrix of similarities generated by T5 was converted into a pairwise dissimilarity matrix using the Kendall rank correlation coefficient. Hierarchical clustering was conducted in R using the AGNES agglomerative clustering (39), and the “flexible” weight average linkage approach, with the *par.method* set to 0.8. The dissimilarity matrix was generated using Pearson correlations, computed using the get_dist() function from the “factoextra” library. The rect.hclust function from R’s “cluster” package was used to cut the hierarchical clustering solution such that the participants were partitioned into 1 through 7 clusters.

### Neuroimaging data acquisition and analysis

Neuroimaging data was available for 66 of the 78 PPA patients. A control sample was used as a reference for quantifying the magnitude of atrophy in our PPA patients. The sample included 24 cognitively normal older adults as control participants, also recruited at MGH (mean age 67.4 ± 4.9; 12 females; mean education 15.7 years ± 2.3). These control participants underwent a neurological and cognitive assessment to confirm the absence of a medical history of neurologic or psychiatric conditions, a structured interview of the participant and an informant by a neurologist, neurological examination, and a neuropsychological test battery (UDS 3.0), and were determined to be clinically normal with the Clinical Dementia Rating scale (CDR=0). All controls had normal brain structure based on MRI and low cerebral amyloid based on quantitative analysis of ^11^C-PiB PET data (FLR DVR < 1.2). MRI data were collected from participants on a Siemens 3-Tesla MAGNETOM Tim Trio scanner using a 12-channel phased-array head coil. Structural MRI data were acquired using a T1-weighted MPRAGE sequence with the following parameters: TR=2530 ms, TE=3.48 ms, flip angle=7 degrees, number of interleaved sagittal slices=176, field of view=256 mm, voxel size=1 mm isotropic. Each participant’s MPRAGE data underwent intensity normalization, skull stripping, and automated segmentation of cerebral white matter to locate the gray-white boundary via FreeSurfer v6.0, which is documented and freely available for download online (http://surfer.nmr.mgh.harvard.edu). Defects in the surface topology were corrected (69), and the gray/white boundary was deformed outward using an algorithm designed to obtain an explicit representation of the pial surface. All cortical surface derivatives were visually inspected for technical accuracy and were manually edited when necessary. Cortical thickness was calculated as the closest distance from the gray/white boundary to the gray/CSF boundary at each vertex on the tessellated surface. For each cluster of patients, we performed a whole-cortex, vertex-wise analysis of cortical thickness compared to controls to identify the spatial topography of atrophy in each patient cluster. For this analysis, we registered all participants’ thickness data to fsaverage space and smoothed them geodesically with full-width-half-maximum (FWHM) of 10 mm. The results of these analyses were inspected via maps of statistical significance at each vertex point overlaid on the average cortical surface template. For these exploratory analyses, a statistical threshold of p < 0.01 was used.

### Linguistic Feature Discovery

#### Extracting language features

The transcribed speech of the patients and healthy controls was analyzed for linguistic features using the Stanza natural language processing toolkit (45).

#### Including high- and low-frequency verbs

In addition to the features automatically extracted from the parser, we separated high- and low-frequency verbs based on the logic discussed in the introduction. We used three different corpora of spoken language, COCA, Switchboard and Santa Barbara. The distribution of verbs produced in these corpora showed a similar discontinuity that distinguished a subset of verbs as highly frequent ones versus other verbs as shown in Supplementary Material 1.

#### Obtaining the residual for each language feature

Counting the number of occurrences for each language feature requires normalization. Such analyses need to control for the overall size of the language sample, as well as basic differences in the relative frequency of certain linguistic features. For example, nouns are produced far more frequently than adjectives or number words in ordinary speech in healthy individuals. Hence, determining whether a particular linguistic feature occurs less often than expected in an individual with PPA requires that the frequency of the linguistic feature be assessed relative to benchmarks established in controls. This allows us to directly assess the degree to which the frequencies differ from normal controls, not just from the other variants.

The number of occurrences of each linguistic feature is reported relative to the frequencies observed in healthy controls. The process of controlling for both language sample size and the base rates of each linguistic feature was achieved by comparing the observed counts against expected counts. A standardized residual is defined as the difference between the observed value and the expected value divided by the square root of the expected value, 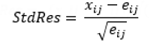. The expected value of the *ij*-th cell in a contingency table, *e_ij_*, is given by multiplying the sum of the counts in the *i*th row (the row marginal total), Σ*x_i_*, by the sum of the counts in the *j*th column (the column marginal total), Σ*x_j_*, and dividing by the total sample size, *N*, 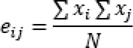. It has been noted that residuals tend to increase with increases in the number of observations (46). The problem of inflated residuals can be addressed by dividing the raw residuals, *x_ij_ − e_ij_*, by the standard error of the raw residuals, 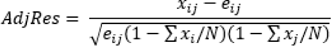 (70), producing an adjusted residual. If the variables associated with *i* and *j* are independent, then the counts across columns will be proportionally the same across the rows. Under these conditions, expected values will closely approximate observed values. However, if the variables are not independent, the proportion of counts across columns will differ across rows. Non-independence will be observed when, for example, the proportion of counts across different linguistic features differs across individuals. The above procedure can be modified to produce residuals showing deviations from healthy controls. This can be accomplished by replacing the column counts across patients with column counts across healthy controls, with the column counts weighted so that the total of the column counts for patients and healthy controls are the same. The column counts would thus indicate the expected proportion of counts across linguistic features based on healthy controls, while the row counts would control for the relative size of the language sample of each patient. The resulting residuals would hence reflect the degree to which a linguistic feature differs from what would be observed from healthy controls. Calculating the residuals for each patient across all of the linguistic features results in a matrix that also specifies residuals for each linguistic feature across all of the patients. The residuals across patients for each linguistic feature can be used to determine which linguistic features are impaired in each PPA variant.

#### Correcting for multiple comparisons

To assess whether linguistic features were in deficit for particular variants of PPA, three dummy variables were created for each variant. For the nfvPPA dummy variable, patients were coded with 1 if they belonged to the nfvPPA-cluster and 0 otherwise. The same approach was taken for lvPPA and svPPA. The classic approach to adjust the significance level for multiple statistical tests is the Bonferroni method, which is α/C, where C is the number of comparisons. This method is widely used for problems with independent multiple comparisons. However, in situations in which the predictors are correlated, the Simes method can be used. In this method, all *C p*-values are sorted from smallest to largest. An adjusted significance level is calculated for each position of the list based on α*k*/C, where α is the alpha level (e.g., 0.05) and *k* is the position in the ordered list. Correcting for multiple comparisons using the Simes method indicated that correlations were significant for *p* < .01278. Table 1 contains all of the negative correlations that met the Simes correction cutoff.

The phylogenetic tree was produced using the *fviz_dend()* function available in the *factoextra* library in R by setting the type argument to “phylogenic”, the k argument to 3, and the phylo_layout argument to “layout.gem”. The fviz_dend() function uses the GEM force-directed layout algorithm (71). In such graphs, the distance between the vertices is less important than how the vertices are connected (72).

#### Predicting PPA variant using language features

The point-biserial correlations demonstrate that a wide range of linguistic features are associated with the different variants of PPA. Predictive modeling was conducted using multinomial logistic regression based on the *multinom* function from the *nnet* package in R. The regression included four categories: nfvPPA, lvPPA, svPPA, and healthy control. The input was the raw counts indicating the number of times a patient produced a particular linguistic feature. The healthy controls served as the baseline reference category in the model.

#### Multinomial logistic regression

To protect against over-fitting, we used 10-fold cross-validation over 100 repetitions. The model’s performance is the average accuracy across the folds and repetitions. Cross-validation was conducted using the trainControl function in the *caret* package in R.

## Data Availability

Codes and data used in this work can be accessed upon sending a request to Dr. Bradford Dickerson at.

## Acknowledgments

We thank the research participants and their care partners, without whose contributions this work would not have been possible. We also thank the Athinoula A. Martinos Center for biomedical imaging support.

## Author contributions

Conceptualization: NR, PW, and BCD; methodology: NR, MB, AT, PW, and BCD; investigation: NR, MQ, DH, MB, AT, BW, PW, and BCD; visualization: NR, MB, AT, PW, and BCD; funding acquisition: NR, PW, and BCD; project administration: NR, BW, MQ, DH, PW, and BCD; writing–original draft: NR, PW, and BCD; writing–review and editing: NR, MQ, DH, MB, AT, BW, PW, and BCD.

