## Supplemental Materials for "Using Generative Artificial Intelligence to Classify Primary Progressive Aphasia from Connected Speech"

**Supplementary Materials**

**Supplementary material 1.**

The verb distribution according to Switchboard Dialog Act Corpus (A) and Santa Barbara (B).

**A**

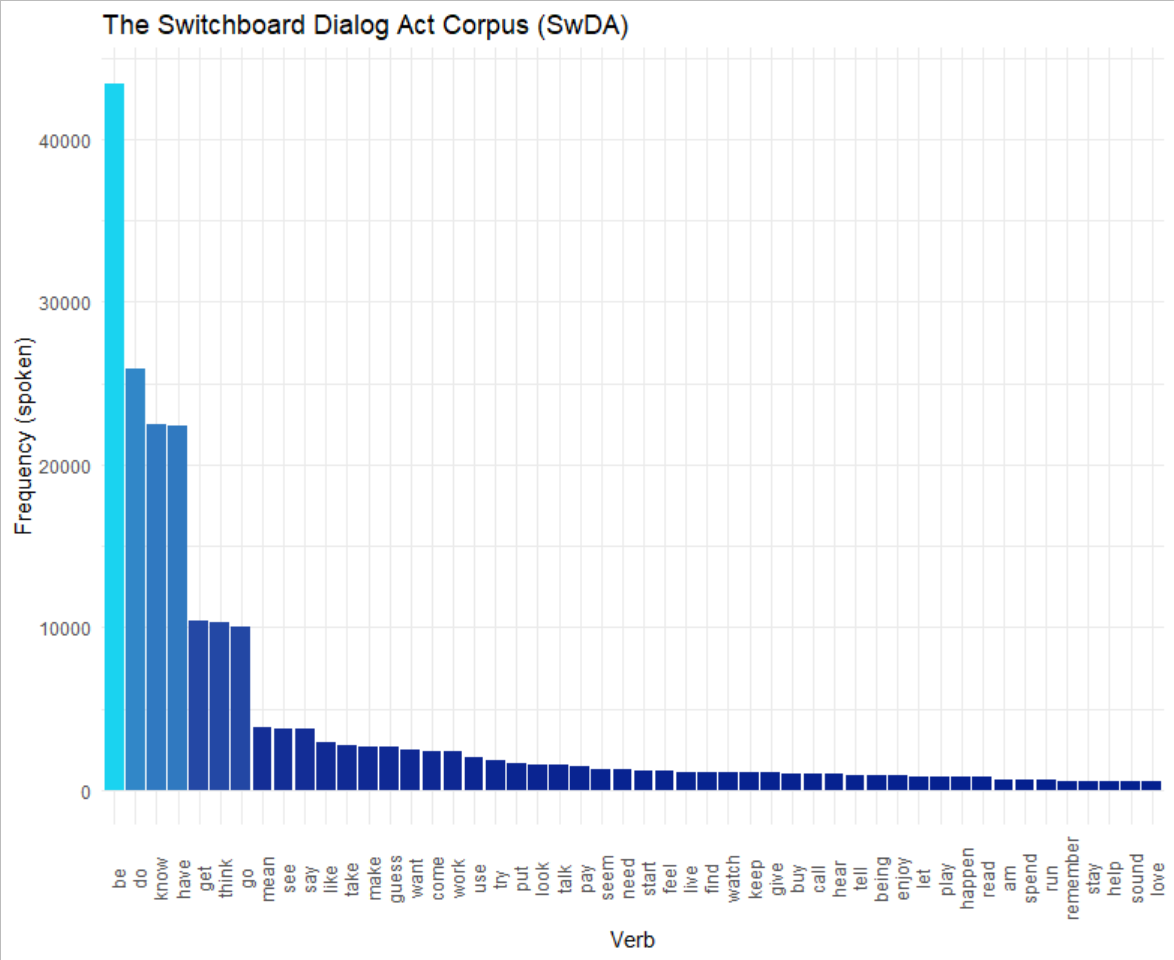

**B**

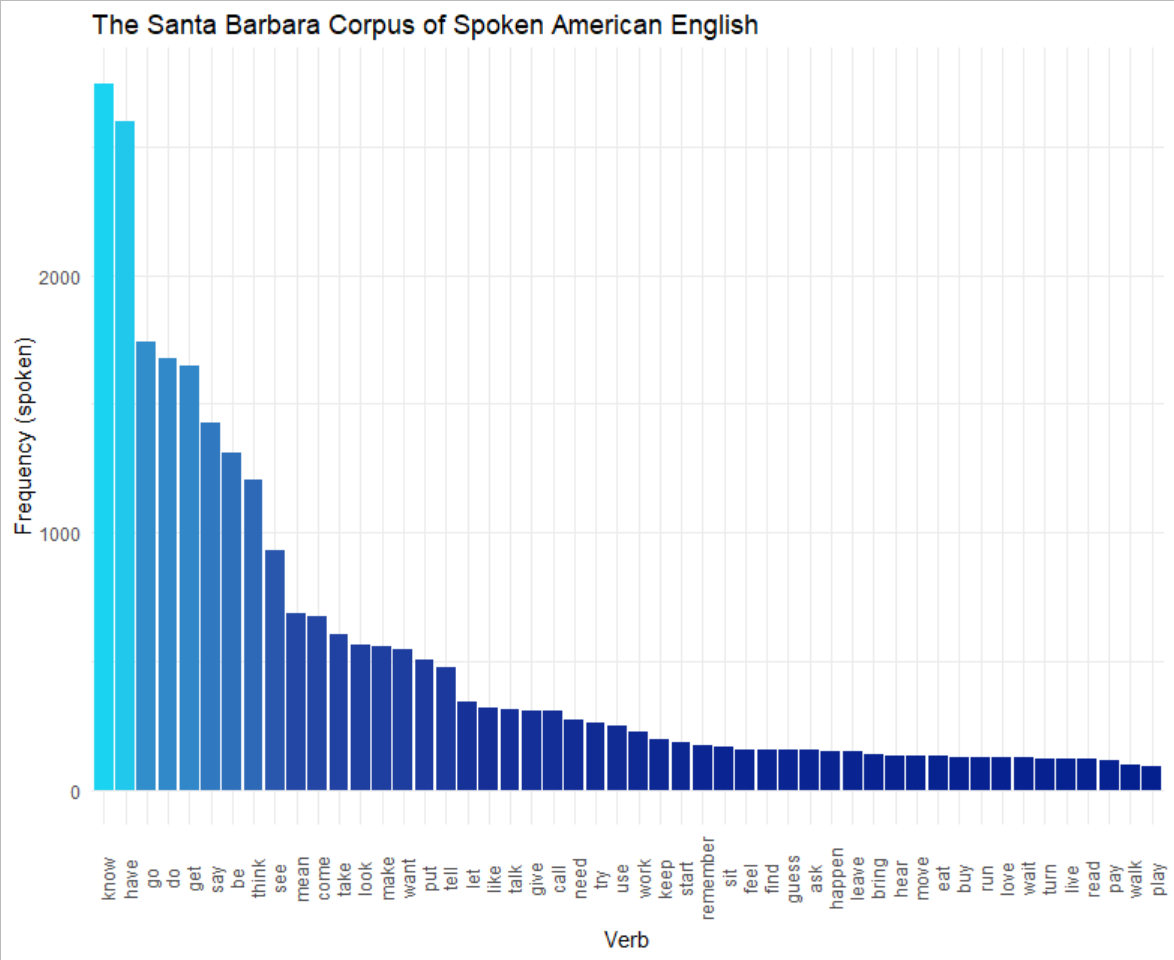

**Supplementary material 2**

The definitions of POS tags and dependency relationships derived from the Stanza automatic parser.

| Art | Articles are determiners which specify definiteness (e.g., the) and indefiniteness (e.g., *a, an*) |
| --- | --- |
| NN (NOUN) | Part of speech for words that function in a sentence as subject, object or prepositional object. Nouns often refer to persons, places, things, animals, or ideas, e.g., *girl*, *kite* |
| Rel | Lexical feature indicating a relative pronoun, determiner, numeral or adverb,, e.g., *The wardrobe that / which leads to Narnia. A fish who is on the dock.* |
| det | Dependency relation that holds between a nominal head and its determiner, e.g., *This task is difficult.* |
| DT | Part of speech that modifies nouns or noun phrases. They may indicate whether the noun is referring to a definite or indefinite element of a class. |
| compound | Dependency relation indicating noun compounds, e.g., *phone book*. |
| nmod | Dependency relation indicating a nominal dependent of another noun or noun phrase that functionally corresponds to an attribute, e.g., *a street across from the beach; boat in the water.* |
| Masc | Pronouns with the lexical feature of grammatically masculine, e.g., *he, him, his*. In the current dataset, these often serve as noun modifiers when used in the possessive, e.g., *The son flew his kite*. |
| Ind | The lexical feature of being non-specific indefinite. It distinguishes something that is specific and known with something that is general and unknown. In English, the feature is carried on definite and indefinite articles, e.g., *A beach. A man and a wife* |
| acl | Dependency relation labeling a clausal modifier of a noun, e.g., *A kid nearby flying a kite*. |
| Def | Lexical feature carried in a determiner specified that something is known and concrete, or unknown and general, e.g., *the kite.* |
| IN | Part of speech typically establishing a spatial (e.g., *in, on, under*), temporal (e.g., *after, during*), subordinating conjunction (e.g., *although, because*) or abstract (e.g., *of, for, via*) relationship. They typically combine with exactly one complement, most often a noun phrase (or determiner phrase). |
| case | The case dependency relation indicates categories of nouns and pronouns with respect to their grammatical function (e.g., subject, object, possession). They are indicated by prepositions (e.g., *house on the street*) and clitics, e.g., *the boy’****s*** *fishing pole*. |
| Sing | The lexical feature Sing typically uses inflectional morphology on nouns to denote one person, animal, or thing, *The son is flying a kite. The house has a big tree in the yard.* |
| VBZ (VERB) | The verb form indicating 3^rd^ person singular present, in which the verb ends with -s or –es, e.g. *Looks like a family singing. There seems to be someone fishing.* |
| VBG (VERB) | The verb form indicating a gerund or present participle. The gerund and present participle are the -ing form of a verb when used as an adjective (e.g., *I am going to remember all this. A little boy flying a kite.* |
| obl | The obl dependency relation uses a preposition to indicate a nominal (noun, pronoun, noun phrase) functions as a non-core (oblique) argument or adjunct, by attaching to a verb, adjective or adverb, e.g., *In the background, there is someone sailing*. *There’s a dog running after the boy.* |
| EX | Part of speech of a nominal that appears in an argument position but does not satisfy any argument (e.g., subject, direct object) of the predicate, e.g., *There is a flag. They’re over there.* |
| Expl | Dependency relation that specifies an expletive, that is, nominals that appear in an argument position of a predicate that do not satisfy any semantic roles of the predicate, e.g., *There is a flag. It is always nice to be near the water.* |
| amod | Dependency relation for adjectival modification, e.g., *The old fellow is catching a fish. a picnic basket on the blanket.* |
| JJ (ADJ) | Adjectives are words that typically modify nouns and specify properties, e.g., *There is a single boat. A little girl is playing.* |
| Ger | Lexical feature specifying a gerund, that is, a verbal noun with the morphological form of the present participle, *The boy’s flying a kite*. *A girl doing something with the stand*. |
| WDT | *Wh*-determiner; e.g., *So those are the main things that I see. The wife is getting something to drink which looks like wine.* |
| P3 | The third person refers to one or more participants that are neither speakers nor addressees, e.g., She’s having fun; *Looks like they are singing*. |
| Pres | The lexical feature specifying the present tense, which denotes actions that are in progress or events that usually happen. |
| NNP (PROPN) | The proper noun, singular, is a noun (or nominal content) that is the name (or part of the name) of a specific individual, place, or object, e.g., *Could be Charlestown*. It is *Memorial day weekend*. *Mom is pouring a glass of wine*. |
| Part | Lexical feature indicating a participal; e.g., present or past participle. In this dataset, mostly present participle  *Dad is reading. He is fishing. They have got a shovel*. |
| VERB_LowFreq | Verbs serving as the root of the sentence not included in the following list of high-frequency verbs: *be, have, do, go, say, know, get* and *think*. |
| nmod: poss | Lexical feature specifying possessive personal pronoun or determiner; e.g., *My glasses; their house;* |
| PRP_poss | Part of speech of possessive pronouns, e.g., His dog is running. *The woman has her shoes off.* |
| advcl | An adverbial clause modifier is a clause which modifies a verb or other predicate, as a modifier not as a core complement; e.g., *dog running behind him trying to catch him*. *She is getting ready for a picnic*. |
| acl:relcl | A relative clause modifier of a nominal dependency relation in which a clause modifies a nominal. The acl:relcl relation points from the head of the modified nominal (e.g., fisherman) to the head of the relative clause (e.g., standing), e.g., *A fisherman who is standing on a pier.* |
| Neut | Pronouns with the lexical feature of grammatically neutral gender, e.g., *it must be a shovel.* |
| MD | The part of speech modal verbs, Modal verbs that take bare infinitives; e.g., *I can see sandals*. *She must be playing in the sand*. |
| TO | Part of speech for the word *to*, specifies the infinitive form, e.g., *I never know what to call it*. |
| xcomp | An open clausal complement of a verb or an adjective, a predicative or clausal complement without its own subject; e.g., *She is getting ready for a picnic*. *The water looks pretty calm*. *A boy trying to pull a kite*. |
| Verb_HighFreq | Verbs serving as the root of the sentence included in the following list of high-frequency verbs: *be, have, do, go, say, know, get* and *think*. |
| mark | Dependency relation tag indicating a subordinate clause, e.g., …after insurgents attacked. *He says that you like to swim. Looks like they’re having a picnic. The father is reading while she’s getting lunch.* |
| Inf | Feature specifying that a verb is in the infinitive (citation) form, e.g., *Can’t think of that; Let’s go back for a minute. I want to think about it.* |
| VB (VERB) | Part of speech for the base form of a verb; e.g., *Let’s start with the kite*. *A car waiting to drive home*. |
| RP (PART) | Part of speech indicating a particle, that is, function word specifying grammatical categories not covered by other parts of speech. In this study, the majority of particles were words that can serve as adpositions that completed phrasal verbs, e.g., *A sailboat out on the lake. Catching up with some of his reading. Cut down the tree.* |
| Acc | A lexical feature specifying that the word serving as the direct object of a verb. In English, accusative case is specified in pronouns (e.g., *me, her, him, them*), and to a lesser extent indicated in prepositions. |
| Int | A lexical feature specifying interrogative determiners (*what, which, where, whose*), which modify a noun or pronoun in a direct or indirect question, e.g., *I know what that is;*  *I don’t know whose house that is; It could be where I live.* In the current dataset, these |
| WP | Part of speech indicating a WH pronoun, e.g., *who, which, whom, what, whose*. In the current corpus, many of the WH pronouns are used with interrogative determiners (Int), e.g., *I don’t know what that is*, but not always, e.g., *Maybe the little boy who is flying the kite are from the house.* |
| Fin | Lexical feature of a verb indicating expression of a mood. Common moods include 1) the indicative (a statement that something has happened), 2) the imperative (an order or request for action), 3) the conditional (a statement that an action would have taken place but actually did not), and 4) the potential (a statement that an action is possible but not certain), 5) the subjunctive (a statement expressing actions that are subjective or otherwise uncertain). |
| ccomp | A dependency relation expressing a clausal complement that is a core argument of a verb or adjective, e.g., *I think we have gone through this. I know what they are. I think he has caught a fish.* |
| Prs | A lexical feature specifying a person or possessive personal pronoun or determiner, e.g., *Looks like she is ready for a picnic. He is flying a kite. It is a seashore with a house in front. A dog is chasing him.* |
| advmod | A dependency relation for an adverb or adverbial phrase that modifies a predicate or a modifier word, e.g., *That is about as much detail as I can make out. The man just caught a fish. He is really relaxed.* |
| PRP (PROPN) | Part of speech for personal pronouns, that is, function words used as substitutes for noun phrases; e.g., *She is building a sandcastle. He caught a fish. They’re reading a book. That’s it.* |
| Nom | A lexical feature of pronouns specifying nominal case, that is, the case indicating that the nominal serves as the subject of a clause, e.g., *Maybe they are listening. It can be anything. She has a bucket.* |
| RB (ADV) | Part of speech for adverbs, which are words that typically modify verbs. They may also modify adjectives and other adverbs (e.g., *very briefly*); e.g., *I always put those in. I am not sure about that. It was a really nice day. Coming back to the picnic.* |
| Dem | Lexical feature for a demonstrative pronoun, determiner, numeral, or adverb which stands in for nouns by pointing to something or someone specific, e.g., *That’s the car. I’m not sure about that. Water is over here. Somebody here fishing.* |
| nmod:poss | A dependency relation indicating a possession, e.g., *Their house is on the water. His shoes are here. Here is somebody’s house. Looks like a family’s picnic.* |

Title case 🡪 Lexical features and Inflectional features

ALL CAPS 🡪 Parts of speech

all lowercase 🡪 Dependency relation

Bold 🡪 base

Underline 🡪 target

**Supplemental material 3**

Normalized confusion matrix for the classification of healthy controls and three PPA variants.

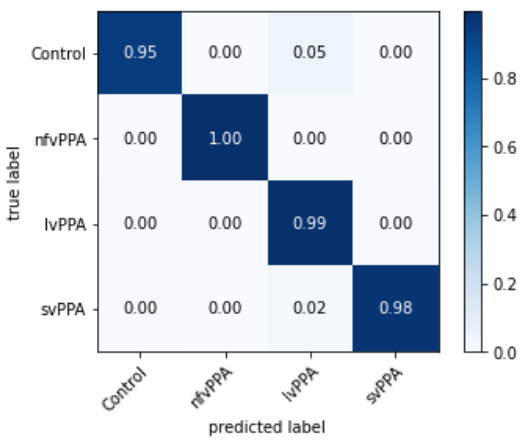
